# Non-Alcoholic Fatty Liver Disease (NAFLD) in Children and Adolescents taking Atypical Antipsychotic Medications: A Protocol for Systematic Review and Meta-Analysis

**DOI:** 10.1101/2021.03.20.20249020

**Authors:** Reem Hatem, Faisal A. Nawaz, Ghadah A. Al-Sharif, Mohammad Almoosa, Wid Kattan, Christos Tzivinikos, Lila Amirali, Ammar Albanna

## Abstract

Atypical Antipsychotic medications are commonly prescribed to children and adolescents and are associated with important adverse effects including weight gain and metabolic syndrome. Non-Alcoholic Fatty Liver Disease (NAFLD) is not only the most common pediatric liver disease, but can also be associated with serious complications including liver cirrhosis. Given that both NAFLD and Atypical Antipsychotics (AAP) are associated with metabolic syndrome, we aimed to comprehensively examine the association between AAP and NAFLD in children and adolescents. We will conduct a systematic review following the PRISMA guidelines, of English-language literature published between 1950 and 2020, exploring NAFLD in subjects younger than 18 years on AAP.

## INTRODUCTION

There is an increasing trend in prescribing atypical antipsychotic medications (AAP) to children and adolescents ^1^, that merit a better understanding of their long-term adverse effects. Metabolic complications such as weight gain, hyperlipidemia and insulin resistance are known side effects of AAP ^2^, and studies have suggested that youth are at an increased risk of these side effects compared to older adults ^3^. It is, therefore, reasonable to speculate that children on AAP may be at a greater risk of disorders associated with metabolic syndrome, including non-alcoholic fatty liver disease (NAFLD).

Pediatric NAFLD is the most common cause of pediatric liver disease, with a prevalence of approximately 9.6% ^4^. The prevalence significantly increases to approximately 77% among obese children ^5^, suggesting its association with metabolic syndrome ^6^. NAFLD comprises a spectrum of liver diseases that range from the accumulation of fat in hepatocytes (steatosis) and inflammatory liver changes (non-alcoholic steatohepatitis <NASH>) to more serious complications such as liver cirrhosis ^7^. The “two-hit” theory attempts to explain the pathogenesis of NAFLD ^8^; the first-hit is caused by the accumulation of triglycerides in hepatocytes causing liver steatosis, and the second-hit is understood to be due to many factors including oxidative stress and increased cytokines leading to inflammatory changes.

Pediatric NAFLD is associated with serious long term adverse outcomes, including increased risk of mortality and increased risk of liver cirrhosis requiring transplantation ^9^.

Multiple risk factors have been linked to the etiology of NAFLD. Obesity has been consistently identified as an important risk factor for pediatric NAFLD ^5^. Insulin resistance is another risk factor for NAFLD; Nobili et al prospectively followed 84 children with NAFLD and reported that they were almost always insulin-resistant regardless of their Body Mass Index (BMI) ^10^. Moreover, sedentary lifestyle and high fructose consumption have been described as risk factors for NAFLD ^11,12^. Furthermore, epidemiological studies identified male sex, older age, and Hispanic ethnicity as risk factors for childhood NAFLD ^4^. Additionally, genetic, cellular and hormonal factors ^13^ have been found to influence the transition to inflammatory hepatic changes.

Liver biopsy is the gold standard diagnostic test for NAFLD^14^, although it is an invasive procedure that can be associated with serious complications. Liver Function Tests (LFT) are amongst first line investigations for NAFLD ^15^. However, the use of LFT to diagnose NAFLD is challenging due to the low sensitivity of this method and discrepancy around appropriate cut-off values ^7^. Moreover, normal LFT level does not exclude the presence of advanced NAFLD ^16^. In contrast, hepatic ultrasonography (US) is a safe, non-invasive and widely available imaging tool for the assessment of NAFLD. In adults, it has acceptable sensitivity and specificity (100% and 90% respectively), especially when the percentage of liver fat exceeds 20 % ^17^. Similar statistical properties were documented in a pediatric study ^18^. Other radiological diagnostic tools for NAFLD include Computed Tomography (CT), Magnetic Resonance Imaging (MRI) and Magnetic Imaging Spectroscopy (MRS) ^19^.

Very little is known regarding the risk of NAFLD in children and adolescents treated with AAP, despite their association with metabolic complications that are considered risk factors for the development of NAFLD.

The objective of our study is to conduct a comprehensive systematic review of the available literature in order to examine the association between AAP use and NAFLD in children and adolescents.

## METHODS

In order to capture all relevant literature on NAFLD among children and adolescents on AAP, we plan to conduct two systematic literature reviews. In our first search, we aim to identify studies that assess for NAFLD in children and adolescents taking AAP. Given that we expect paucity in such studies based on a pilot search performed, we will include studies of different designs including cohort and case-control studies, case reports and case series. In our second systematic search, we will comprehensively review AAP trials in children and adolescents, attempting to identify any reports of NAFLD, whether this was documented as a primary outcome, secondary outcome, or as an incidental finding in these studies.

Our systematic review will be performed according to this predefined protocol that describes the objectives, search strategy, eligibility criteria and evaluation methods according to the PRISMA (Preferred Reporting items for Systematic Reviews and Meta-Analysis) guidelines ^20^.

### Systematic review methodology

We will conduct two systematic literature reviews. The search is restricted to English-literature from January 1, 1950 until March 31, 2021. 1950 was chosen as the starting date for the literature search as it coincides with the development of the first antipsychotic.

1. In our first search, we aim to identify studies that assess for NAFLD in children and adolescents taking AAP. Given that we expect paucity in such studies based on a pilot search, we will include studies of different design including cohort and case-control studies, case reports and case series. Terms: “second generation neuroleptics”,” antipsychotics”, “neuroleptics”, generic and brand names: see table 1. Also, the following variations of Pediatric NAFLD: “NAFLD”, “NASH”, “nonalcoholic fatty liver disease”, “nonalcoholic steatohepatitis”, “hepatic steatosis”, “fatty liver disease”, “nonalcoholic fatty liver”, “fatty liver”. In our second search, we will include the following keywords: “Atypical antipsychotics”, “atypical neuroleptics”.
2. In our second systematic search, we will comprehensively review AAP trials in children and adolescents, attempting to identify any reports of NAFLD, whether this was documented as a primary outcome, secondary outcome, or as an incidental finding during these studies. The following databases will be used to conduct the search: MEDLINE, OVID, Cochrane, and Cumulative Index to Nursing and Allied Health Literature CINAHL. We also searched the following databases up to 2016: Embase, Web of Science, BIOSIS Previews and PsycINFO.

A systematic, computer-assisted search of the following databases will be performed: Ovid Medline, Embase, Web of Science, BIOSIS Previews, Psychinfo, Cumulative Index to Nursing and Allied Health Literature CINAHL, will be conducted by three medical students R.A., F.N., G.A., M.A. at Mohammed Bin Rashid University (MBRU), and the search strategy will be supported by MBRU librarian Shakeel Tegginmani at MBRU Library. Furthermore, the bibliographies of retrieved and relevant articles will be manually searched for relevant articles. Full English-language articles published in peer-reviewed journals will be included in this review.

**Table 1.**
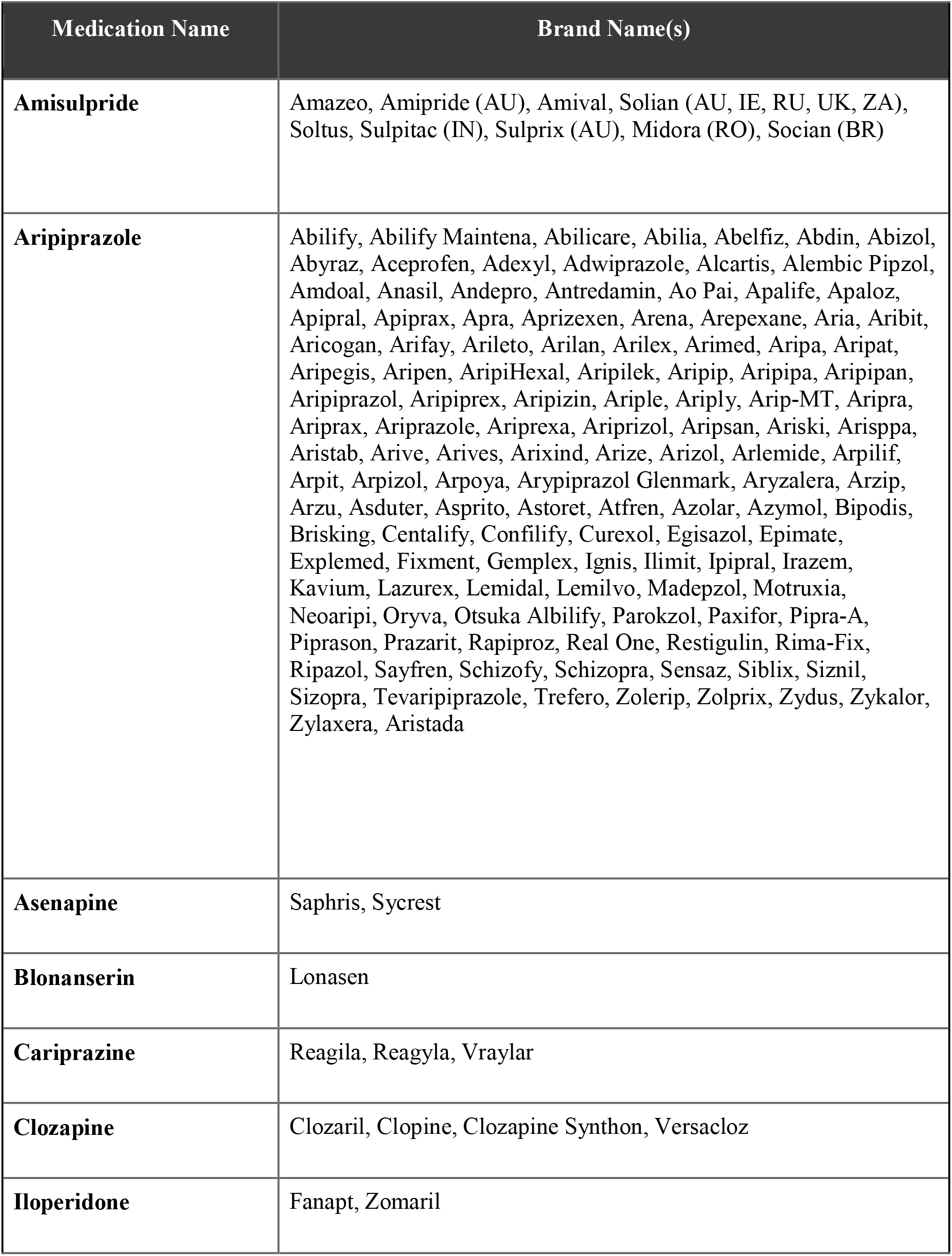
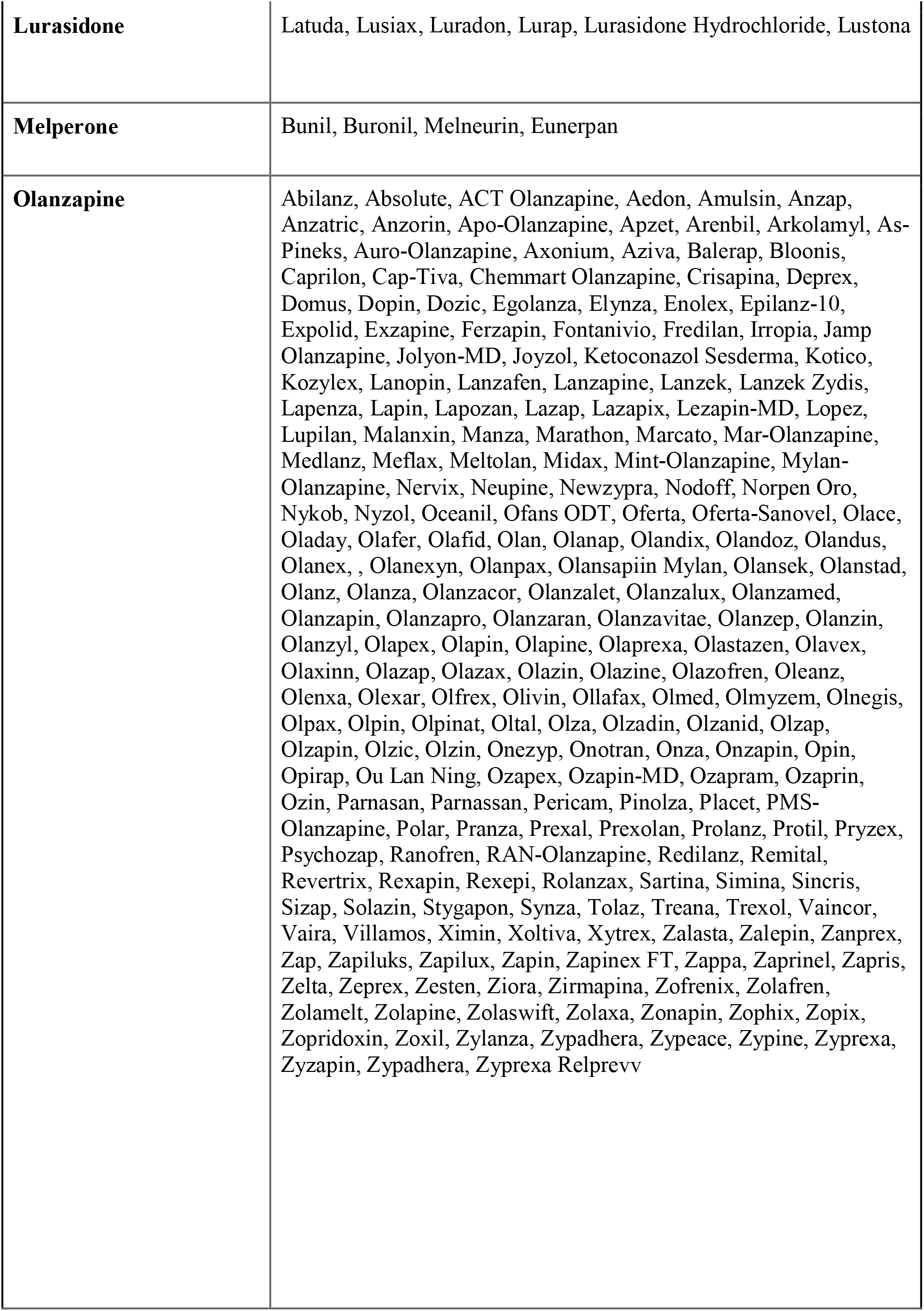
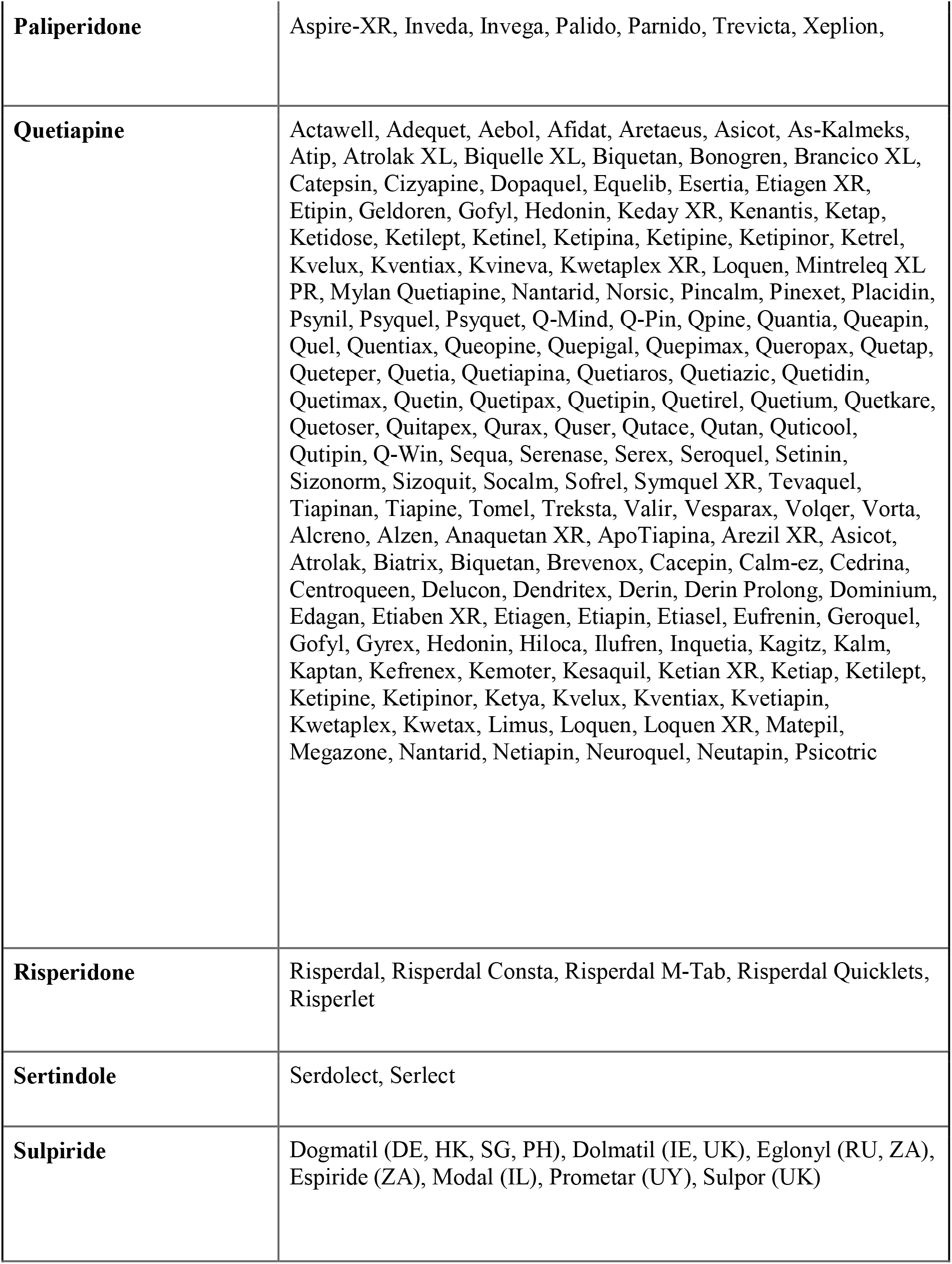
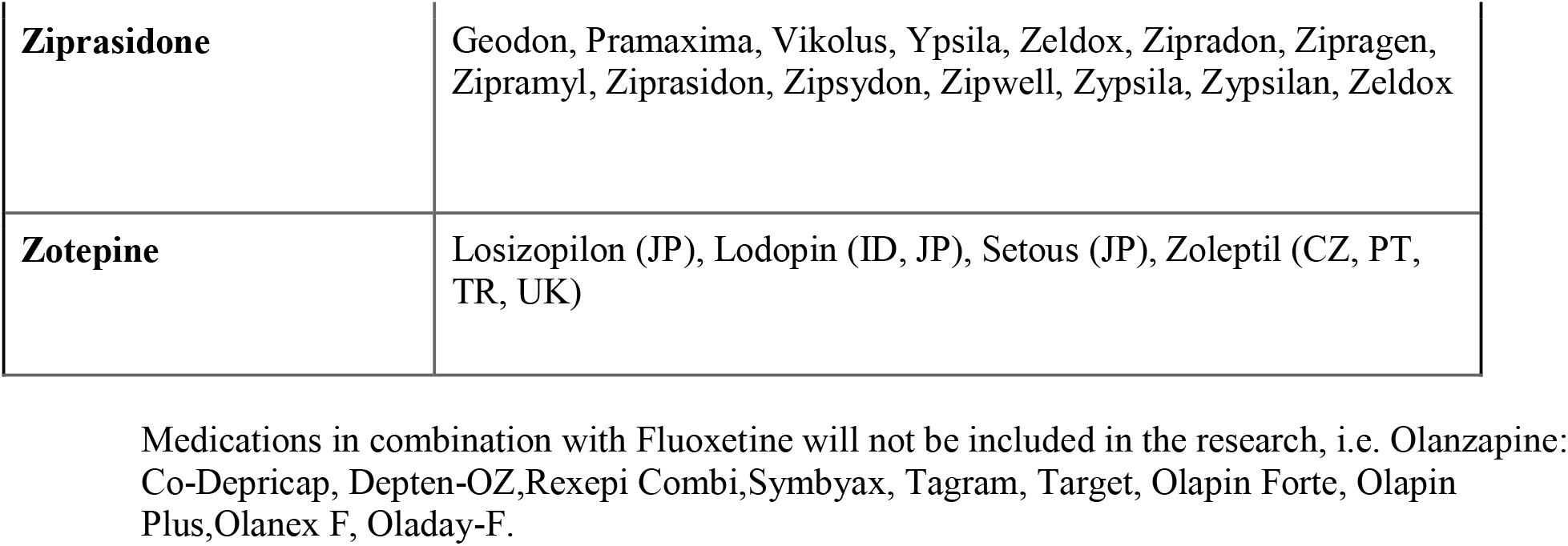

As mentioned earlier, we will conduct two searches that will be limited to children and adolescents younger than 18 years of age. The first search will include the following variations of AAP: “atypical antipsychotics”, “atypical neuroleptics”,” second generation antipsychotics”, “second generation neuroleptics”,” antipsychotics”, “neuroleptics”, and for AAP:

### Eligibility Criteria

We determined the following inclusion and exclusion criteria based on a literature review and the objectives of this study. All English-Language published medication trials of AAP in subjects younger than 18 years of age that reported NAFLD as an outcome, as assessed by radiological method (including liver ultrasound) or liver biopsy, will be included in this review. Moreover, prospective and retrospective observational studies of children and adolescents on AAP with reported NAFLD indicators will be included. Finally, case series and reports of NAFLD in children and adolescents on AAP will be included. We will exclude conference abstracts, editorials, letters to editors, treatment guidelines, and studies published as abstracts only. Reviews (both systematic and non-systematic) will be also excluded, however, the bibliographies of relevant papers will be reviewed. We will exclude studies in which AAP were used as an add-on or in combination with other medications (such as antiepileptics, FGAs SSRIs) and medications that are a combination of AAP and SSRI including : (Fluoxetine), but we will include studies where AAP constitute one arm of the study. We will also exclude studies examining patients with pre-existing medical conditions including eating disorders. Our search will be limited to studies in English language and non-human studies will be excluded. The review and comparison of results will be conducted using Endnote.

### Outcomes

The primary outcome of the present study is NAFLD in children and adolescents, as assessed by either liver biopsy or using a radiological tool including hepatic ultrasound, MRI, MRS, and Transient Elastography (TE) among children and adolescents on AAP. Despite the relationship between NAFLD and metabolic syndrome, we did not include variables of metabolic syndrome in our outcomes given that it is not specifically the focus of our review and that it has been previously studied, including a recent meta-analysis ^21^. Secondary outcome will include changes in Liver Enzymes.

### Data-Extraction

Three investigators, R.A., F.N., G.A. and M.A., will independently review titles and abstracts of retrieved studies and exclude duplicates and irrelevant studies based on the eligibility criteria above. All of the potentially eligible abstracts will be further assessed for eligibility by thoroughly reviewing their full-texts. Results will be compared and discrepancies will be resolved by consensus and by consulting the investigators, A.A., C.T. and L.A., when needed. Cohen’s Kappa Coefficient will be calculated as a measure of inter-rater agreement. All studies that meet the eligibility criteria will be included in our analyses. Data extraction will be carried out using a standard form.

Quality of the studies will be assessed using: STROBE^(34)^ for case cohort, case-control and cross-sectional studies, Newcastle-Ottawa Scale for non-randomized studies, and the quality of RCTs will be reported using Grading of Recommendations, Assessment, Development and Evaluations GRADE method.

If the results allow for conducting a meta-analysis, we will report the outcomes by calculating the weighted pooled estimate of changes in the outcomes. The risk of bias will be evaluated using three types of homogeneity tests: 1) forest plot, 2) Cochrane’s Q test (chi-squared), and 3) Higgins I2 statistics. In the forest plot, as shown, with greater overlap between the confidence intervals indicating greater homogeneity.

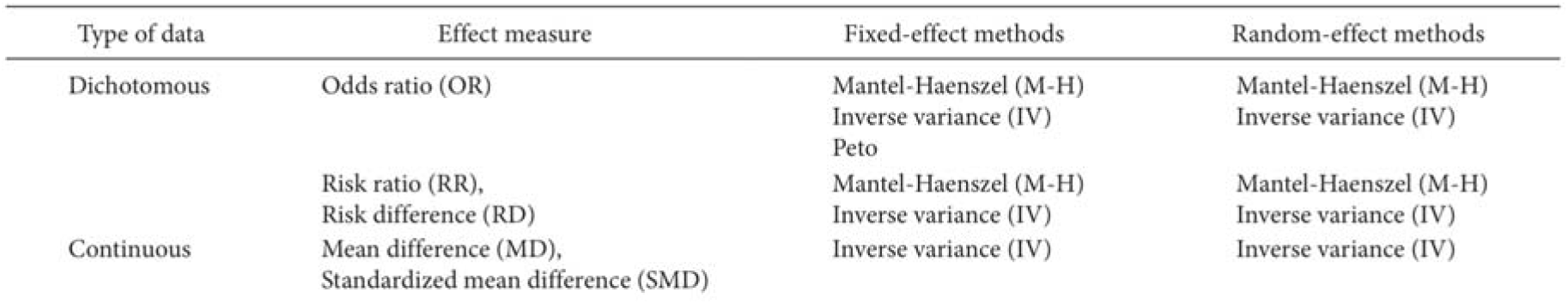

**Figure (35)**

## RESULTS and CLINICAL SIGNIFICANCE

The search results will be presented using the PRISMA flowchart. This study will comprehensively review the literature for NAFLD in children and adolescents taking atypical antipsychotics. This is specifically important due to the current trends in overprescribing AAP in children below the age of 18 ^(36)^. The results of this study may inform clinical guidelines for AAP use in children and adolescents.

## Data Availability

All available data mentioned in the manuscript has been referenced as per the relevant research guidelines.

## Notes

### Competing Interest Statement

The authors have declared no competing interest.

### Funding Statement

No external funding was received by any of the authors for this project

### Author Declarations

*INSERT IRB STATEMENT*

## REFERENCES

1. Pringsheim, T., Lam, D. and Patten, S. The Pharmacoepidemiology of Antipsychotic Medications for Canadian Children and Adolescents: 2005–2009. Journal of Child and Adolescent Psychopharmacology. 2011; 21 (6): 537–543. DOI: 10.1089/cap.2010.0145. PMID: 22136092

2. Fedorowicz, V. and Fombonne, E. Metabolic side effects of atypical antipsychotics in children: a literature review. Journal of Psychopharmacology. 2005; 19 (5): 533–550. DOI: 10.1177/0269881105056543. PMID: 16166191

3. Hammerman, A., Dreiher, J., Klang, S., Munitz, H., Cohen, A. and Goldfracht, M. Antipsychotics and Diabetes: An Age-Related Association. Annals of Pharmacotherapy. 2008; 42 (9): 1316–1322. DOI: 10.1345/aph.1L015. PMID: 18664607

4. Schwimmer, J., Deutsch, R., Kahen, T., Lavine, J., Stanley, C. and Behling, C. Prevalence of Fatty Liver in Children and Adolescents. PEDIATRICS. 2006; 118 (4): 1388–1393. DOI: 10.1542/peds.2006-1212. PMID: 17015527

5. Chan, D., Li, A., Chu, W., Chan, M., Wong, E., Liu, E., Chan, I., Yin, J., Lam, C., Fok, T. and Nelson, E. Hepatic steatosis in obese Chinese children. International Journal of Obesity. 2004; 28 (10): 1257–1263. DOI: 10.1038/sj.ijo.0802734. PMID: 15278103

6. Fu, J., Non-alcoholic fatty liver disease: An early mediator predicting metabolic syndrome in obese children? World Journal of Gastroenterology. 2011; 17 (6): 735. DOI: 10.3748/wjg.v17.i6.735. PMID: 21390143

7. Vajro, P., Lenta, S., Socha, P., Dhawan, A., McKiernan, P., Baumann, U., Durmaz, O., Lacaille, F., McLin, V. and Nobili, V. Diagnosis of Nonalcoholic Fatty Liver Disease in Children and Adolescents. Journal of Pediatric Gastroenterology and Nutrition. 2012; 54 (5): 700–713. DOI: 10.1097/MPG.0b013e318252a13f. PMID: 22395188

8. AlKhater, S. Paediatric non-alcoholic fatty liver disease: an overview. Obesity Reviews. 2015; 16 (5): 393–405. DOI: 10.1111/obr.12271. PMID: 25753407

9. Bhala, N., Angulo, P., van der Poorten, D., Lee, E., Hui, J., Saracco, G., Adams, L., Charatcharoenwitthaya, P., Topping, J., Bugianesi, E., Day, C. and George, J. The natural history of nonalcoholic fatty liver disease with advanced fibrosis or cirrhosis: An international collaborative study. Hepatology. 2011; 54 (4): 1208–1216. DOI: 10.1002/hep.24491. PMID: 21688282

10. Nobili, V., Marcellini, M., Devito, R., Ciampalini, P., Piemonte, F., Comparcola, D., Sartorelli, M. and Angulo, P. NAFLD in children: A prospective clinical-pathological study and effect of lifestyle advice. Hepatology. 2006; 44 (2): 458–465. DOI: 10.1002/hep.21262. PMID: 16871574

11. Nseir, W., Hellou, E. and Assy, N. Role of diet and lifestyle changes in nonalcoholic fatty liver disease. World J Gastroenterol. 2014; 20 (28): 9338–9344. DOI: 10.3748/wjg.v20.i28.9338. PMID: 25071328

12. Jin, R. et al. Dietary fructose reduction improves markers of cardiovascular disease risk in Hispanic-American adolescents with NAFLD. Nutrients. 2014; 6 (8): 3187–3201. DOI: 10.3390/nu6083187. PMID:25111123

13. Hassan, K., Bhalla, V., Regal, El M. E. & A-Kader, H. H. Nonalcoholic fatty liver disease: a comprehensive review of a growing epidemic. World J. Gastroenterol. 2014; 20 (34): 12082– 12101. DOI:10.3748/wjg.v20.i34.12082. PMID: 25232245.

14. Kleiner, D. E. et al. Design and validation of a histological scoring system for nonalcoholic fatty liver disease. Hepatology.2005; 41 (6): 1313–1321. DOI: 10.1002/hep.20701. PMID: 15915461

15. Mann, J. P., Goonetilleke, R., and McKiernan, P. Paediatric non-alcoholic fatty liver disease: a practical overview for non-specialists. Archives of Disease in Childhood. 2014; 100(7):673–677. DOI:10.1136/archdischild-2014-307985. PMID: 25633064

16. Manco, M. et al. Metabolic syndrome and liver histology in paediatric non-alcoholic steatohepatitis. Int J Obes. 2007; 32 (2). 381–387.

17. Dasarathy, S. et al. Validity of real time ultrasound in the diagnosis of hepatic steatosis: a prospective study. Journal of Hepatology 51, 1061–1067 (2009). DOI: 10.1038/sj.ijo.0803711. PMID: 18087267

18. Shannon, A. et al. Ultrasonographic quantitative estimation of hepatic steatosis in children with Nonalcoholic Fatty Liver Disease (NAFLD). Journal of Pediatric Gastroenterology and Nutrition. 2011; 53 (2):190–195. DOI:10.1097/MPG.0b013e31821b4b61. PMID: 21788761.

19. Lee, S. S. and Park, S. H. Radiologic evaluation of nonalcoholic fatty liver disease. World J. Gastroenterol. 2014; 20 (23): 7392–7402. DOI: 10.3748/wjg.v20.i23.7392. PMID: 24966609.

20. Liberati, A. et al. The PRISMA Statement for Reporting Systematic Reviews and Meta-analyses of Studies That Evaluate Healthcare Interventions: Explanation and Elaboration. J Clin Epidemiol. 2009; 62, (10): e1–34. DOI: 10.1016/j.jclinepi.2009.06.006. PMID: 19631507.

21. Almandil, N. B. et al. Weight Gain and Other Metabolic Adverse Effects Associated with Atypical Antipsychotic Treatment of Children and Adolescents: A Systematic Review and Meta-analysis. Pediatr Drugs. 2013; 15 (2): 139–150. DOI: 10.1007/s40272-013-0016-6. PMID: 23519708

22. Holtmann, M., Kopf, D., Mayer, M., Bechtinger, E. & Schmidt, M. H. Risperidone-associated Steatohepatitis and Excessive Weight-Gain. Pharmacopsychiatry. 2003; 36 (5): 206–207. DOI: 10.1055/s-2003-43045. PMID: 14571356.

23. Kumra, S., Herion, D., Jacobsen, L. K., Briguglia, C. & Grothe, D. Case Study: Risperidone-Induced Hepatotoxicity in Pediatric Patients. Journal of the American Academy of Child & Adolescent Psychiatry. 1997; 36 (5): 701–705. DOI: 10.1097/00004583-199705000-00022. PMID: 9136506.

24. Copur, M. & Erdogan, A. Risperidone rechallenge for marked liver function test abnormalities in an autistic child. Recent Pat Endocr Metab Immune Drug Discov. 2011; 5 (3): 237–239. DOI: 10.2174/187221411797265872. PMID: 21913889.

25. Lui, S. Y., Tso, S., Lam, M. and Cheung, E. F. C. Possible olanzapine-induced hepatotoxicity in a young Chinese patient. Hong Kong Med. 2009; 15 (5): 394–396. DOI: PMID: 19801701.

26. Erdogan, A. et al. Risperidone and liver function tests in children and adolescents: A short-term prospective study. Prog. Neuro-Psychopharmacol. Biol. Psychiatry. 2008; 32 (3): 849–857. DOI: 10.1016/j.pnpbp.2007.12.032. PMID: 18258348.

27. Erdogan, A. et al. Six Months of Treatment with Risperidone May Be Associated with Nonsignificant Abnormalities of Liver Function Tests in Children and Adolescents: A Longitudinal, Observational Study from Turkey. Journal of Child and Adolescent Psychopharmacology. 2010; 20 (5): 407–413. DOI: 10.1089/cap.2009.0113. PMID: 20973711.

28. Karaman, M. G. et al. Liver function tests in children and adolescents receiving risperidone treatment for a year: A longitudinal, observational study from Turkey a. Int J Psych Clin Pract. 2011; 15 (3): 204–208. DOI: 10.3109/13651501.2011.582537. PMID: 22121930.

29. Pérez-Iglesias, R. et al. Effect of antipsychotics on peptides involved in energy balance in drug-naive psychotic patients after 1 year of treatment. J Clin Psychopharmacol. 2008; 28 (3): 289– 295. DOI: 10.1097/JCP.0b013e318172b8e6. PMID: 18480685.

30. Reynolds, G. P. & Kirk, S. L. Metabolic side effects of antipsychotic drug treatment--pharmacological mechanisms. Pharmacology and Therapeutics. 2010; 125 (1): 169–179. DOI: 10.1016/j.pharmthera.2009.10.010. PMID: 19931306.

31. Bonhaus, D. W. et al. RS-102221: a novel high affinity and selective, 5-HT2C receptor antagonist. NP. 1997; 36 (4-5): 621–629. DOI: 10.1016/s0028-3908(97)00049-x. PMID: 9225287.

32. Oh, K. J. et al. Atypical antipsychotic drugs perturb AMPK-dependent regulation of hepatic lipid metabolism. AJP: Endocrinology and Metabolism. 2011; 300 (4): E624–E632. DOI: 10.1152/ajpendo.00502.2010. PMID: 21224484.

33. Lauressergues, E. et al. Antipsychotic drug action on SREBPs-related lipogenesis and cholesterogenesis in primary rat hepatocytes. Naunyn-Schmied Arch Pharmacol. 2010; 381 (5): 427–439. DOI: 10.1007/S00210-010-0499-4. PMID: 20333360.

34. da Costa B, Cevallos M, Altman D, Rutjes A, Egger M. Uses and misuses of the STROBE statement: bibliographic study. BMJ Open. 2011; 1 (1): e000048–e000048. DOI: 10.1136/bmjopen-2010-000048. PMID: 22021739

35. Ahn E, Kang H. Introduction to systematic review and meta-analysis. Korean J Anesthesiol. 2018; 71 (2): 103–112. DOI: 10.4097/kjae.2018.71.2.103. PMID: 29619782

36. Harrison J, Cluxton-Keller F, Gross D. Antipsychotic Medication Prescribing Trends in Children and Adolescents. Journal of Pediatric Health Care. 2012;26(2):139–145. DOI: 10.1016/j.pedhc.2011.10.009. PMID: 22360933

